# Performance of a Protein Language Model for Variant Annotation in Cardiac Disease

**DOI:** 10.1101/2024.06.04.24308460

**Authors:** Aviram Hochstadt, Chirag Barbhaiya, Anthony Aizer, Scott Bernstein, Marina Cerrone, Leonid Garber, Douglas Holmes, Robert J Knotts, Alex Kushnir, Jacob Martin, David Park, Michael Spinelli, Felix Yang, Larry A Chinitz, Lior Jankelson

**Affiliations:** NYU Langone Health and the NYU Grossman School of Medicine

## Abstract

**Introduction:** Genetic testing is a cornerstone in the assessment of many cardiac diseases. However, variants are frequently classified as Variants of Unknown Significance (VUS), limiting the utility of testing.

Recently, the DeepMind group (Google, USA) developed AlphaMissense, a unique Artificial Intelligence (AI) based model, based on language model principles for the prediction of missense variant pathogenicity.

**Objective:** To report on the performance of AlphaMissense, accessed by VarCardio, an open web-based variant annotation engine, in a real-world cardiovascular genetics center.

**Methods:** All genetic variants from an inherited arrhythmia program were examined using AlphaMissense via VarCard.io and compared to the ClinVar variant classification system, as well as another variant classification platform (Franklin by Genoox). The mutation reclassification rate and genotype phenotype concordance were examined for all variants in the study.

**Results:** We included 266 patients with heritable cardiac diseases, harboring 339 missense variants. Of those, 230 (67.8%) were classified by ClinVar as either VUS or non-classified. Using VarCard.io, 198 VUSs (86.1%, CI 80.9-90.3%) were reclassified to either Likely Pathgenic (LP) or Likely benign (LB). The reclassification rate was significantly higher for VarCard.io than for Franklin (86.1% vs 34.8%, p<0.001).

Genotype-Phenotype concordance was highly aligned using VarCard.io predictions, at 95.9% (CI 92.8-97.9%) concordance rate. For 109 variants classified as Pathogenic, LP, Benign or LB by ClinVar, concordance with VarCard.io was high (90.5%).

**Conclusion:** AlphaMissense, accessed via VarCard.io, may be a highly efficient tool for cardiac genetic variant interpretation. The engine’s notable performance in assessing variants that are classified as VUS in ClinVar, demonstrates its potential to enhance cardiac genetic testing.

## Introduction

Genetic testing for monogenic disease has transformed the care of patients affected by a wide spectrum of cardiovascular conditions by enabling the identification of specific mutations which explain individual phenotypes^1^. The enhanced precision that comes with genetic analysis results in improved inference of risk, tailoring of gene-specific targeted therapy (i.e. Nadolol in LQT1), and upstream prevention using cascade screening in family members. In recent cohorts, as many as 50% of patients with genetically linked cardiovascular conditions are found to be positive for a reportable genetic variant.^1–3^ A major limitation of genetic testing remains the finding of a Variant of Unknown Significance (VUS), occurring in up to 40% of genetic tests.^4–7^ Recently, the DeepMind group (Google, USA) developed AlphaFold and AlphaMissense ^8,9^, Artificial Intelligence (AI) based models for the prediction of protein folding and variant pathogenicity, respectively. Here, we report the performance of AlphaMissense, interfaced via VarCard.io^10^, an open web-based variant annotation engine allowing cDNA and protein change queries aligned with Clingen derived gene-disease correlation, and its performance in a real-world cardiovascular genetics center.

## Methods

### AlphaMissesne and VarCard.io

The development and testing of AlphaMissesne have been reported elsewhere^9^. Briefly, AlphaMissense predicts the pathogenicity of all possible amino acid (AA) changes in all known protein sequences. The model is trained by using an unsupervised learning approach, implementing a language model architecture. In an initial pretraining step, deep complex representations of protein sequence data are built by masking and unmasking amino acids at random along a given sequence. By training the model to predict the masked AAs, the model learns the fundamental properties of the human proteome and the compatibility of any AA in any position. In the second step, the model is fine-tuned on a set of variants which are labeled based on either being highly frequent or completely absent from human and primate populations. To allow easy interaction with AlphaMissense predictions, we built VarCard.io, a web engine that allows query of all missense variants by gene name and cDNA or AA change and extracts the AlphaMissense annotation of each variant as either Likely Benign (LB), Likely Pathogenic (LP) or Ambiguous. In addition, VarCard.io extracts the gene-disease correlation for the queried gene, as assessed by ClinGen.

### Patients and annotations

All patients with missense variants found in probands presenting to the NYU Inherited Arrhythmia program were included. All missense variants found in each patient were included. We compared the annotation of AlphaMissense via VarCard.io to the ClinVar^11^ database, as well as to the annotation of an independent commercial classification engine, Franklin^12^ (Genoox, Palo Alto, CA, USA). ClinVar annotations were recorded as per ACMG guidelines to be either Pathogenic (P), Likely pathogenic (LP), Variant of unknown significance (VUS), Benign (B) or likely benign (LB). If the variant did not appear on the ClinVar database it was treated in this study as a VUS unless otherwise specified. If a variant had a mixed classification (i.e. more than one entry in ClinVar), it was treated as a VUS if at least one annotation was a VUS.

### Assessing clinical Genotype-Phenotype relationship

All genes in the database were assessed using the ClinGen^13^ framework (ClinGen, National Institutes of Health) as to their known gene-disease correlation. Patient records were reviewed, and the clinical phenotype was assessed by using all clinical data available including patient’s history, ECG tracings, provocative tests and imaging (e.g. echocardiography, cardiac MRI, Calcium Pyrophosphate imaging). The genotype-phenotype correlation was considered as positively concordant If: 1) a patient had a variant in a gene with known gene-disease correlation compatible with the designated clinical phenotype for this gene by ClinGen, and 2) the variant was classified as P/LP (positive concordance). Alternatively, negative concordance was determined if: 1) a variant was found in a gene not compatible with the patients’ clinical phenotype by ClinGen, and 2) the variant was classified as B/LB. Otherwise, the genotype-phenotype was considered as discordant (i.e. a patient with a P/LP variant in a gene not compatible with the patients’ phenotype). Patients were excluded from analysis if their phenotype was indeterminable due to missing data. Patients included in the study provided written informed consent before inclusion which was approved by the NYU Langone Health IRB committee in accordance with the Helsinki Declaration.

### Statistical analysis

Discrete variables are reported as numbers and rates, 95% confidence intervals are calculated using the Clopper-Pearson method. The statistical significance of rate differences was assessed using the Chi-Square test. Results were considered significant when p-values were <0.05. All calculations were done using R version 3.3.2 (R Foundation for Statistical Computing, Vienna, Austria).

## Results

We reviewed 623 probands in our original database, of which 266 patients were identified as meeting the inclusion criteria of having at least one genetic variant that was classified as a missense variant in a gene recognized to be associated with a cardiac phenotype by ClinGen (Figure-1). These patients had a total of 339 different genetic variants. The most common phenotype was Long QT syndrome, followed by Non-ischemic Cardiomyopathy and Hypertrophic Cardiomyopathy (Figure-1). The most frequently involved genes were SCN5A (implicated in Brugada syndrome, dilated cardiomyopathy and LQT syndrome), followed by KCNQ1, KCNH2 and RyR2 (Figure-2).

**Figure-1.**
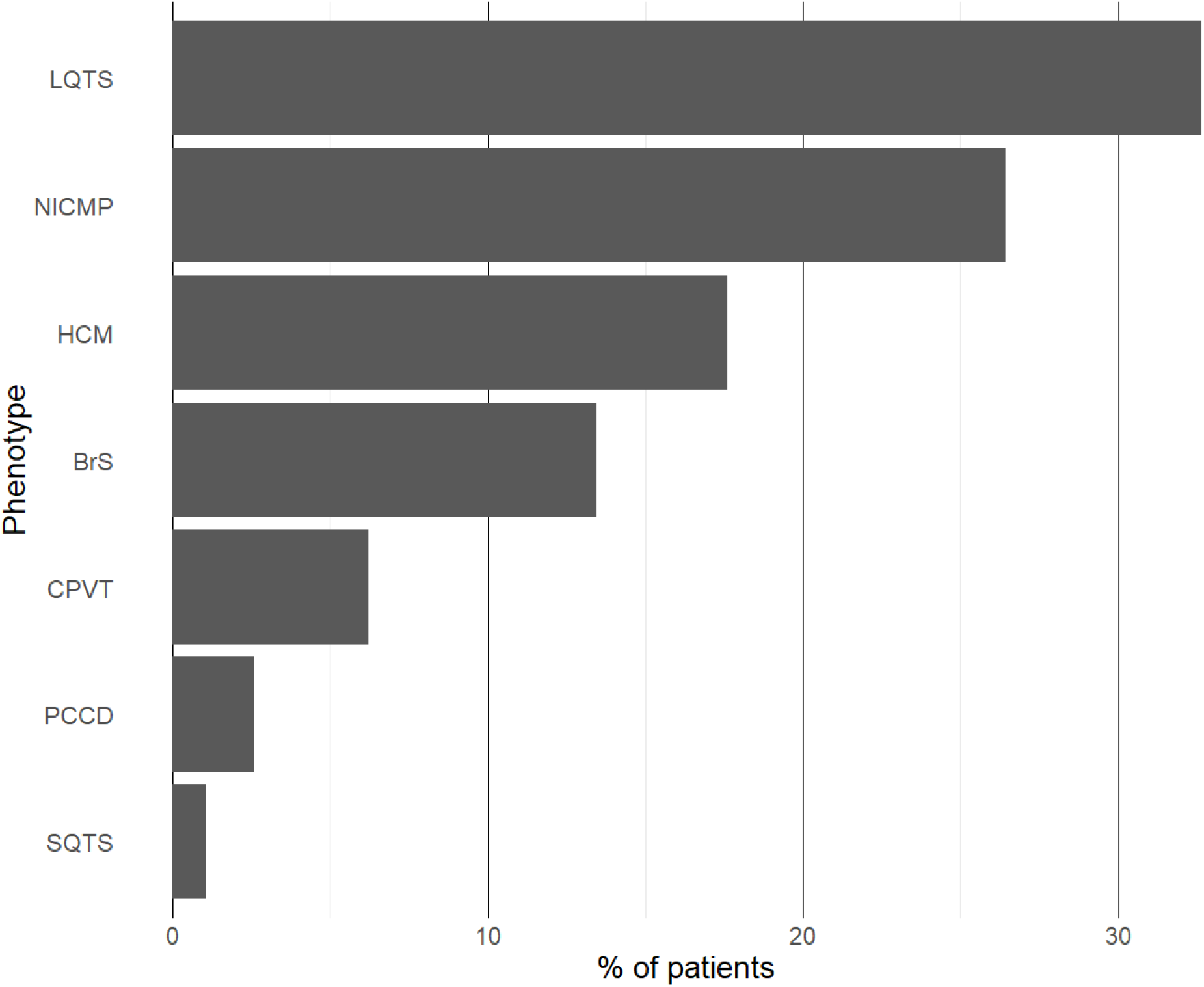
Frequency graph of patients’ most frequent phenotypes in the cohort. LQTS- Long QT syndrome, NICMP- Non-Ischemic Cardiomyopathy, HCM- Hypertrophic Cardiomyopathy. BrS-Brugada Syndrome. CPVT - Catecholaminergic Polymorphic Ventricular Tachycardia. PCCD- Progressive Cardiac Conduction Defect, SQTS- Short QT Syndrome.

**Figure-2:**
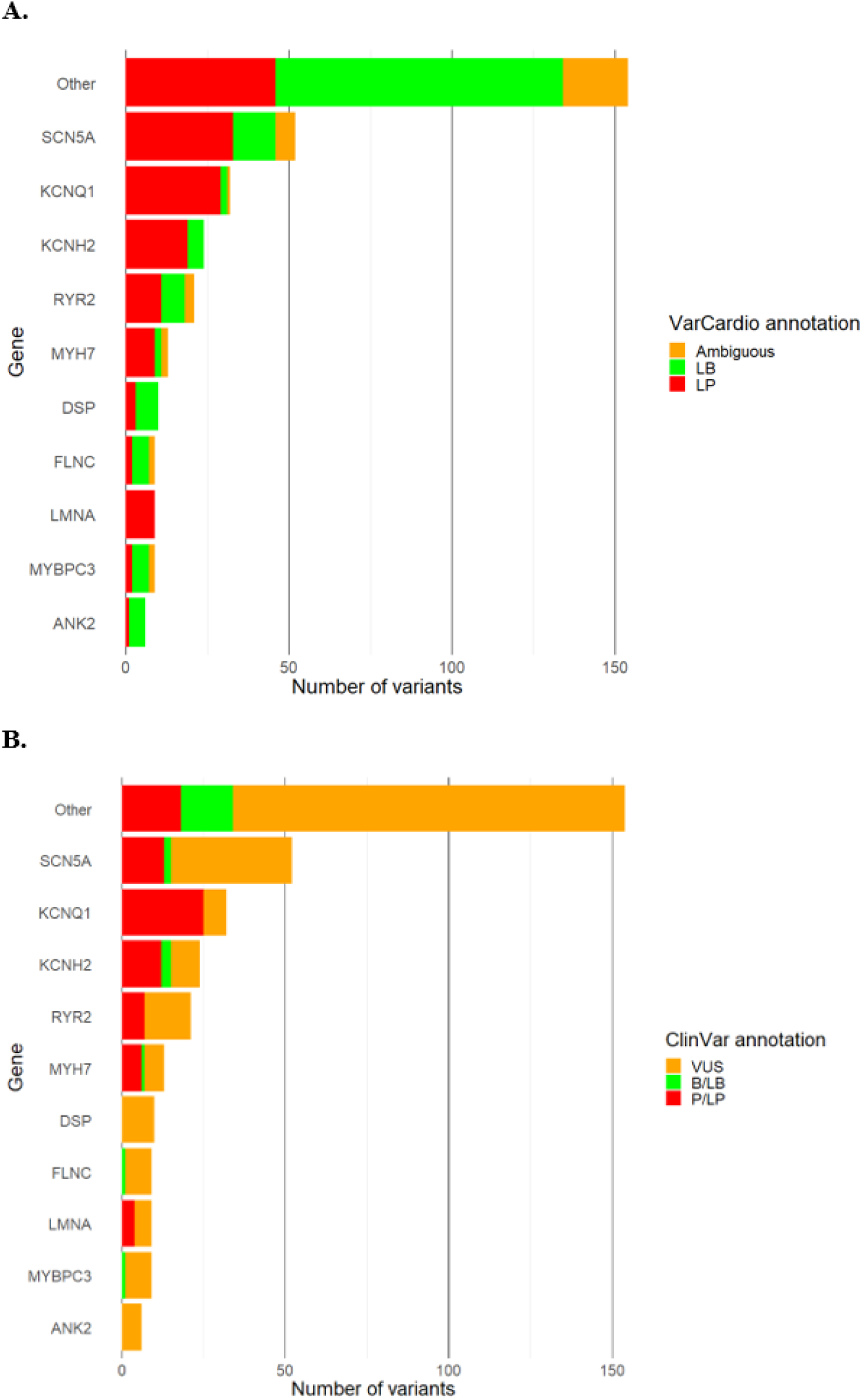
Variant frequency and annotation according to A. VarCard.io, B. ClinVar. P- Pathogenic, LP- Likely Pathogenic, B-Benign, LB- Likely Benign, VUS- Variant of Unknown Significance.

Of the 339 variants tested in VarCard.io, 328 variants had ClinVar entries, of which 110 (33.5%) variants had a Clinically Significant Variant (CSV) annotation of pathogenicity (i.e. P, LP, B or LB without VUS entries), and the rest were classified as VUS or had mixed classification (Table-1).

**Table-1:**
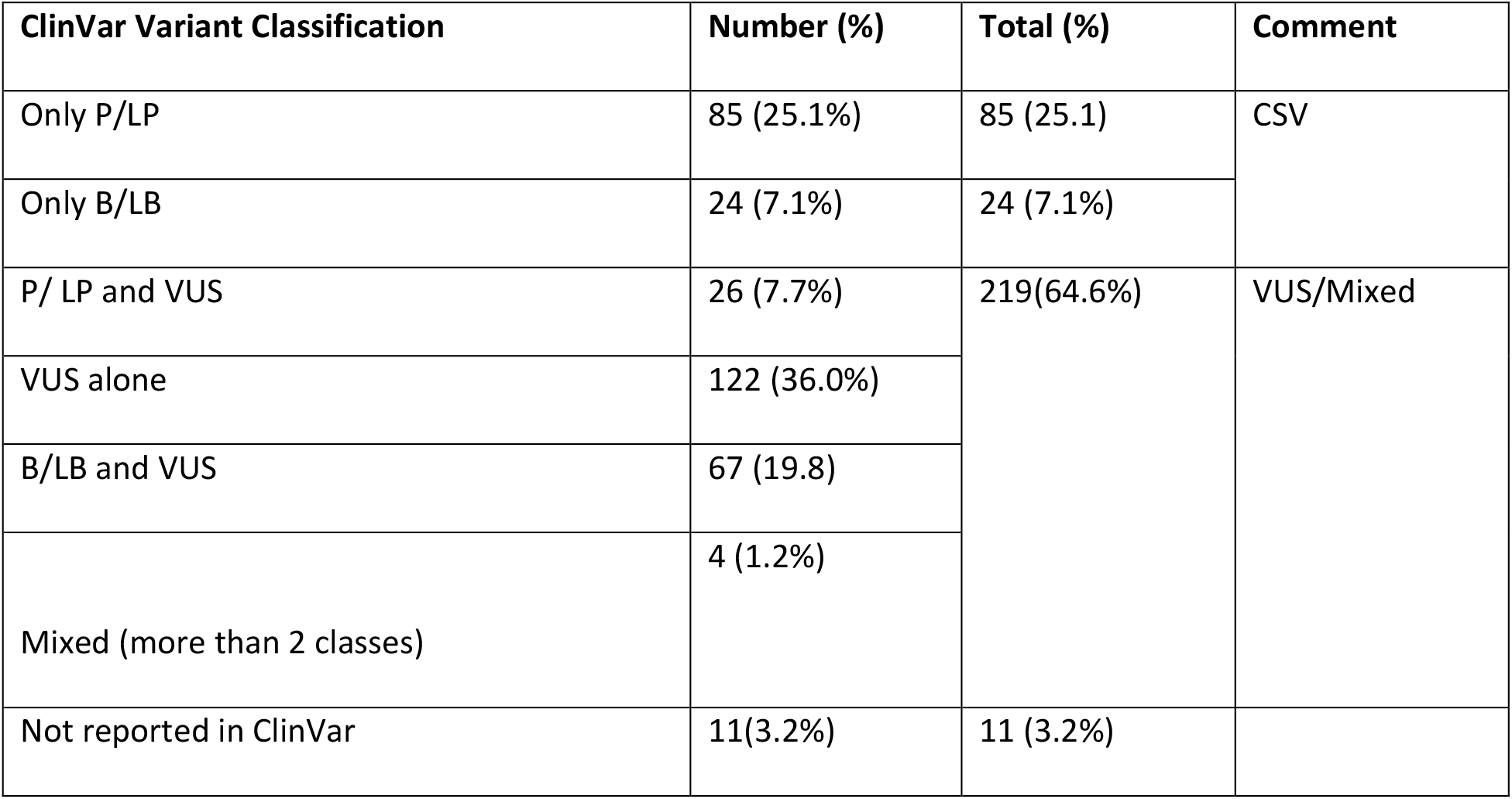
Variant classification according to ClinVar. P- Pathogenic, LP- Likely Pathogenic, B-Benign, LB- Likely Benign, CSV- Clinically Significant Variant, VUS- Variant of Unknown Significance.

| ClinVar Variant Classification | Number (%) | Total (%) | Comment |
| --- | --- | --- | --- |
| Only P/LP | 85 (25.1%) | 85 (25.1) | CSV |
| Only B/LB | 24 (7.1%) | 24 (7.1%) |  |
| P/ LP and VUS | 26 (7.7%) | 219(64.6%) | VUS/Mixed |
| VUS alone | 122 (36.0%) |  |  |
| B/LB and VUS | 67 (19.8) |  |  |
| Mixed (more than 2 classes) | 4 (1.2%) |  |  |
| Not reported in ClinVar | 11(3.2%) | 11 (3.2%) |  |

### Comparison of ClinVar annotation to VarCard.io

For the 230 (67.8%) VUSs (i.e. variants with at least one ClinVar entry as VUS or not reported), 198 variants were reclassified by VarCard.io as either LP or LB, yielding a reclassification rate of 86.1% (CI 80.9-90.3%). These reclassifications were to LB in 111 variants (56.1%, CI 48.8% - 63.1%) and to LP in 87 variants (43.9%, CI 36.9% - 51.2%, Figure-3). Only 32 variants (13.9%) had an ambiguous annotation by VarCard.io.

**Figure-3:**
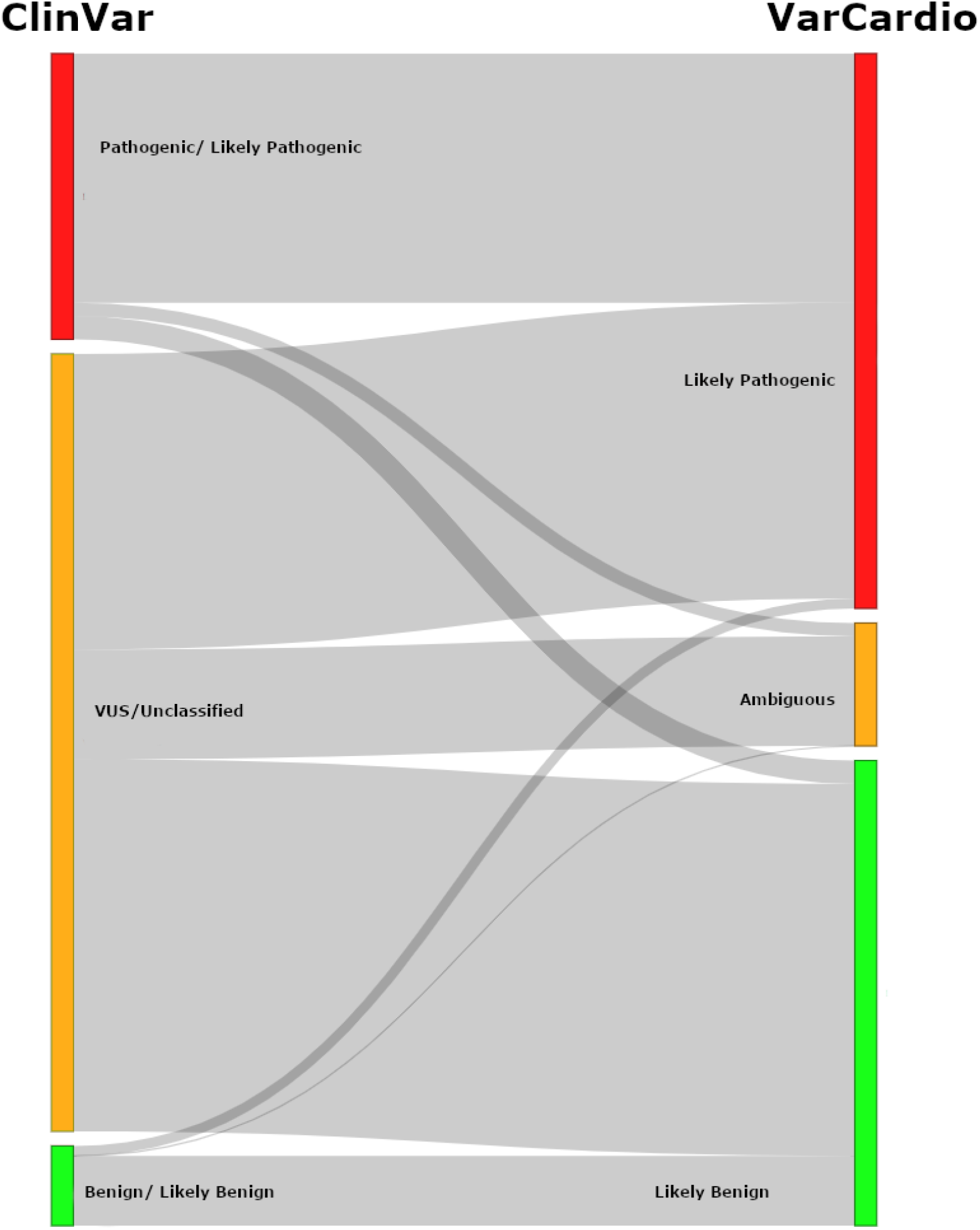
Sankey Diagram showing reclassification of Variant reclassification changes from ClinVar to VarCard.io. VUS – Variant of unknown significance.

Of the 109 variants with a CSV ClinVar annotation (i.e. variants without any VUS entry), 105 remained a CSV, and the other four (3.7 %) were annotated as Ambiguous by VarCard.io. The overall agreement rate between ClinVar and VarCard.io on CSVs (i.e. same annotation in both systems) was 90.5% (CI 83.2-95.3%). This agreement rate was similar for 81 variants with ClinVar P/PL annotation and for 24 variants with ClinVar B/LB annotation (91.4% and 87.5%, respectively, p=0.865).

### VarCard.io genotype-phenotype concordance

Of the 303 variants with a CSV annotation by VarCard.io, 269 variants were related to a patient with a defined phenotype. Of these, 258 variants (95.9%, CI 92.8-97.9%) had concordant genotype-phonotype prediction: 138 (96.5%, CI 92-98.9%) with positive concordance and 120 (95.2%, CI 89.9-98.2%) with negative concordance. There were 34 (11.2%) variants in patients without sufficient clinical data to establish a defined phenotype and thus were not included in this analysis.

### Comparison with another publicly available variant classification engine (Franklin)

Using Franklin on the 230 VUS variants appearing in ClinVar resulted in reclassification to either P/LP or B/LB in only 80 variants, giving a reclassification rate of 34.8% (CI 28.6-41.3%), significantly lower than the 86.1% (CI 80.9%-90.3%) VarCard.io reclassification rate (p<0.001). For the 100 variants with CSV annotation in both ClinVar and Franklin, agreement was 100% (CI 96.4-100).

## Discussion

In this work, we aimed to evaluate the performance of AlphaMissense, interfaced through VarCard.io, an open searchable web-based engine for variant pathogenicity prediction, on a real-world cohort of patients with heritable arrhythmia and cardiomyopathy syndromes (Central illustration). Our main findings are:

1. Using VarCard.io, we reclassify 86.1% of VUSs to either likely pathogenic or likely benign classes. This reclassification rate was significantly greater than the reclassification rate of 35% observed in a widely used commercial variant annotation engine (Franklin).
2. Assigned annotations by VarCard.io were highly concordant with the observed clinical phenotypes, with 95.9% genotype-phenotype concordance.
3. We found a 90.5% agreement rate between ClinVar and VarCard.io on variants with pathogenic or benign annotations.

A diagnosis of VUS in a gene with cardiac manifestations, occurring in up to 40% of genetic tests^4–7^, is a pressing clinical problem which may have direct implications on patients’ risk assessment, specific therapy, possible preventive strategies (such as in PKP2 mutation carriers), family screening opportunities, and patients’ anxiety.^14,15^ As genetic panels become increasingly available and encompass more genes, the incidence of VUSs is also increasing, stressing the need for reliable and accessible variant annotation strategies.

The AlphaMissense variant pathogenicity model has the potential to facilitate the field of genetic testing by significantly enhancing the performance of computational VUS reclassification. The use of AlphaMissense was previously shown to result in the classification of 88.8% of 69.5 million variants present on gnomAD ^9^ to LP or LB, compatible with our finding of 86.1% reclassification rate for ClinVar-reported VUSs.

The major difference in architecture between AlphaMissesne and prevailing popular computational models such as SIFT^16^ and REVEL^17^ is that these models are largely based on labels of pathogenicity guided by sequence homology between species and prevalence in population databases. This introduces inherent biases, as some relatively rare variants are pathogenic, and many cardiac conditions are characterized by age-dependent penetrance. In addition, traditional supervised AI models are often trained on human-provided labels, thereby preserving a circular logic and bias.

The high performance achieved by AlphaMissense is the result of the model’s unique self-supervised architecture, built and trained based on the principals of an LLM. In the case of a language model, each word is represented by a vector (embedding) which captures semantic relationships between words, allowing the model to reason about language, i.e. predict the compatibility of any word in any given context. In the case of AlphaMissense and variant prediction, vectors represent Amino Acids, and their compatibility in each genomic position is computed based on complex properties that capture their “context” within the protein. To further enhance performance, the model is then fine-tuned on a defined set of variants with well-established annotations based on extreme population frequency properties.

While our results demonstrate VarCard.io’s utility, it is important to acknowledge certain limiting aspects. As AlphaMissense is an AI model with predictions based on complex, convoluted representations, there is no simple way to derive linear mechanistic explanations for the different predictions and their pathogenesis in the clinical phenotype. In addition, AlphaMissense does not infer on penetrance nor predict how a variant would interact with a patient’s specific multi-omic inventory to create the specific phenotype (i.e. would not predict if a specific SCN5A variant would cause Brugada Syndrome or Long QT Syndrome). Furthermore, assessing clinical concordance of genes and phenotypes might be biased as both a gene’s function and possibly pathogenicity and a patient’s phenotype may be complex and not straightforward to assess for this study’s purposes. Variants may also have incomplete penetrance and age-related penetrance, thus a patient not having a phenotype at a specific time point does not necessarily mean a specific variant is not pathogenic in nature. Finally, as suggested by ACMG, computational predictions are one component in a matrix of criteria recommended for the clinical evaluation of genetic variants. Thus, although the PP3 criterion was recently upgraded from to “supporting” to “moderate” or “strong”^18^, it should still be considered within the context of other features.

In conclusion, despite the above limitations, VarCard.io, propelled by AlphaMissense, may be a highly efficient tool for cardiac genetic variant interpretation. The AlphaMissense notable performance in assessing variants that are classified as VUS in ClinVar demonstrates the potential of AI as an important step forward in the field of clinical cardiovascular genetics.

## Data Availability

Data will be made available for researchers after IRB approval for the proposed use of the data.

## Abbreviations

VUS: Variant of Unknown Significance
AI: Artificial Intelligence
AA: Amino Acids
LB: Likely Benign
LP: Likely Pathogenic
B: Benign
P: Pathogenic
CSV: Clinically Significant Variant
LQTS: Long QT syndrome
NICMP: Non-Ischemic Cardiomyopathy
HCM: Hypertrophic Cardiomyopathy
BrS: Brugada Syndrome
CPVT: Catecholaminergic Polymorphic Ventricular Tachycardia
PCCD: Progressive Cardiac Conduction Defect
SQTS: Short QT Syndrome.

## Central illustration

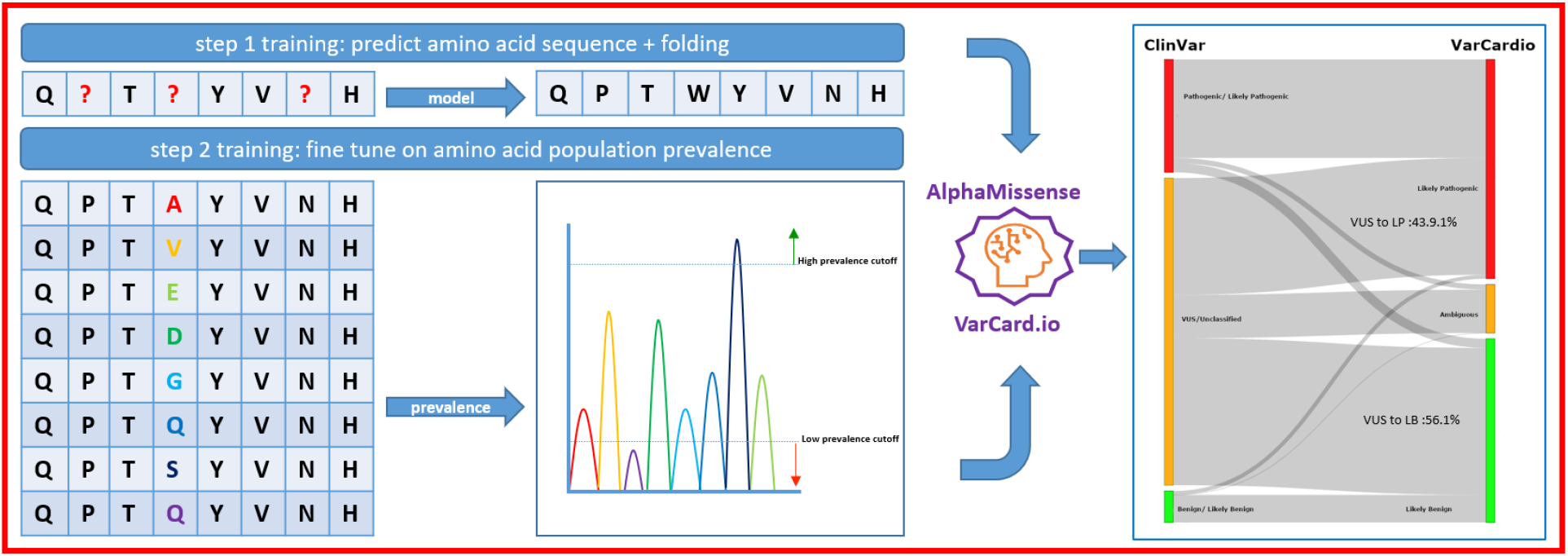

### Central Illustration Figure legend

Architecture and performance of AlphaMissense and VarCard.io. Letters represent amino acid symbols. The AlphaMissense model is built based on principals of a large language model (LLM), aimed at predicting text. In the first step of training, the model is learning to predict amino acid sequences in a self-supervised manner. This process results in the learning of embeddings, i.e. representations of amino acids conditioned on their context (sequence), which are in turn used to infer on protein folding and amino acid compatibility. Next, the model is fine-tuned on a small subset of protein variants with known pathogenicity based on their population frequencies (i.e. extremely frequent and extremely rare). We then use AlphaMissesnse via the VarCard.io search engine to assess variant annotation in a cohort of patients with inherited arrhythmia and cardiomyopathy. LB-VUS-Variant of unknown significance. LB - Likely Benign, LP Likely Pathogenic.

